# REACT-1 round 11 report: low prevalence of SARS-CoV-2 infection in the community prior to the third step of the English roadmap out of lockdown

**DOI:** 10.1101/2021.05.13.21257144

**Authors:** Steven Riley, David Haw, Caroline E. Walters, Haowei Wang, Oliver Eales, Kylie E. C. Ainslie, Christina Atchison, Claudio Fronterre, Peter J. Diggle, Andrew J. Page, Alexander J. Trotter, Thanh Le Viet, Nabil-Fareed Alikhan, Justin O’Grady, The COVID-19 Genomics UK (COG-UK) Consortium, Deborah Ashby, Christl A. Donnelly, Graham Cooke, Wendy Barclay, Helen Ward, Ara Darzi, Paul Elliott

**Affiliations:** School of Public Health, Imperial College London, UK; MRC Centre for Global infectious Disease Analysis and Abdul Latif Jameel Institute for Disease and Emergency Analytics, Imperial College London, UK; Centre for Infectious Disease Control, National Institute for Public Health and the Environment, Bilthoven, The Netherlands; CHICAS, Lancaster Medical School, Lancaster University, UK and Health Data Research, UK; Quadram Institute, Norwich, UK; Department of Statistics, University of Oxford, UK; Department of Infectious Disease, Imperial College London, UK; Imperial College Healthcare NHS Trust, UK; National Institute for Health Research Imperial Biomedical Research Centre, UK; Institute of Global Health Innovation at Imperial College London, UK; MRC Centre for Environment and Health, School of Public Health, Imperial College London, UK; Health Data Research (HDR) UK London at Imperial College; UK Dementia Research Institute at Imperial College

## Abstract

**Background:** National epidemic dynamics of SARS-CoV-2 infections are being driven by: the degree of recent indoor mixing (both social and workplace), vaccine coverage, intrinsic properties of the circulating lineages, and prior history of infection (via natural immunity). In England, infections, hospitalisations and deaths fell during the first two steps of the “roadmap” for exiting the third national lockdown. The third step of the roadmap in England takes place on 17 May 2021.

**Methods:** We report the most recent findings on community infections from the REal-time Assessment of Community Transmission-1 (REACT-1) study in which a swab is obtained from a representative cross-sectional sample of the population in England and tested using PCR. Round 11 of REACT-1 commenced self-administered swab-collection on 15 April 2021 and completed collections on 3 May 2021. We compare the results of REACT-1 round 11 to round 10, in which swabs were collected from 11 to 30 March 2021.

**Results:** Between rounds 10 and 11, prevalence of swab-positivity dropped by 50% in England from 0.20% (0.17%, 0.23%) to 0.10% (0.08%, 0.13%), with a corresponding R estimate of 0.90 (0.87, 0.94). Rates of swab-positivity fell in the 55 to 64 year old group from 0.17% (0.12%, 0.25%) in round 10 to 0.06% (0.04%, 0.11%) in round 11. Prevalence in round 11 was higher in the 25 to 34 year old group at 0.21% (0.12%, 0.38%) than in the 55 to 64 year olds and also higher in participants of Asian ethnicity at 0.31% (0.16%, 0.60%) compared with white participants at 0.09% (0.07%, 0.11%). Based on sequence data for positive samples for which a lineage could be identified, we estimate that 92.3% (75.9%, 97.9%, n=24) of infections were from the B.1.1.7 lineage compared to 7.7% (2.1%, 24.1%, n=2) from the B.1.617.2 lineage. Both samples from the B.1.617.2 lineage were detected in London from participants not reporting travel in the previous two weeks. Also, allowing for suitable lag periods, the prior close alignment between prevalence of infections and hospitalisations and deaths nationally has diverged.

**Discussion:** We observed marked reductions in prevalence from March to April and early May 2021 in England reflecting the success of the vaccination programme and despite easing of restrictions during lockdown. However, there is potential upwards pressure on prevalence from the further easing of lockdown regulations and presence of the B.1.617.2 lineage. If prevalence rises in the coming weeks, policy-makers will need to assess the possible impact on hospitalisations and deaths. In addition, consideration should be given to other health and economic impacts if increased levels of community transmission occur.

## Introduction

National patterns of COVID-19 cases show high temporal variation [1], likely driven by: the degree of recent indoor mixing (both social and workplace), vaccine coverage, intrinsic properties of the circulating lineages, and prior history of infection (via natural immunity). For example, India is experiencing a large wave of cases and deaths coinciding with the emergence of the B.1.617 lineage, after a period of relatively high levels of social mixing with low current levels of vaccination [2]. Meanwhile in Israel, although restrictions on social mixing have largely been relaxed, with high levels of vaccination, case incidence is at low levels, likely driven both by direct and indirect effects of the vaccine [3].

Since 6 January 2021 with the implementation of the third national lockdown in England, regulations to reduce social mixing have been in place, with their relaxation in two stages from 8 March and 12 April as part of a “roadmap”. During the same period, a rapid national vaccination programme, in which individuals were prioritised based on their risk of death if infected, has succeeded in vaccinating 35.5 million people (67.6% of all adults in the UK) with at least one dose (up to 10 May) [4]. Despite the predominant lineage during this period (B.1.1.7) being more transmissible than the prior wild type [5], incidence of hospitalisations and deaths have fallen considerably and are now at only 2.6% and 0.7% respectively of their peak January values (most recent 7-day averages, as reported 10 May [4]).

Surveillance of SARS-CoV-2 in the community through self-swabbing of a representative sample of the population coupled with PCR assays has helped to identify patterns in the epidemic in England that could not be seen reliably with routine testing for cases, hospitalisations and deaths [6]. Here, we report the latest results from the REal-time Assessment of Community Transmission-1 (REACT-1) study [7]. Round 11 of REACT-1 commenced self-administered swab-collection on 15 April 2021 and completed collections on 3 May 2021. We compare the results of REACT-1 round 11 to round 10, in which swabs were collected from 11 to 30 March 2021.

## Results

In round 11 we found 115 positives from 127,408 swabs giving an unweighted prevalence of 0.09% (0.07%, 0.11%) and a weighted prevalence of 0.10% (0.08%, 0.13%) (Table 1). This represents a 50% reduction from round 10 in which unweighted prevalence was 0.16% (0.14%, 0.18%) and weighted prevalence was 0.20% (0.17%, 0.23%).

**Table 1.** Unweighted and weighted prevalence of swab-positivity across nine rounds of REACT-1 and round 11.

| Round | Tested swabs | Positive swabs | Unweighted prevalence (95% CI) | Weighted prevalence (95% CI) | First sample | Last sample |
| --- | --- | --- | --- | --- | --- | --- |
| 1 | 120,620 | 159 | 0.13% (0.11%, 0.15%) | 0.16% (0.13%, 0.19%) | 1/5/2020 | 1/6/2020 |
| 2 | 159,199 | 123 | 0.077% (0.065%, 0.092%) | 0.088% (0.068%, 0.11%) | 19/6/2020 | 7/7/2020 |
| 3 | 162,821 | 54 | 0.033% (0.025%, 0.043%) | 0.040% (0.027%, 0.053%) | 24/7/2020 | 11/8/2020 |
| 4 | 154,325 | 137 | 0.089% (0.075%, 0.11%) | 0.13% (0.096%, 0.15%) | 20/8/2020 | 8/9/2020 |
| 5 | 174,949 | 824 | 0.47% (0.44%, 0.50%) | 0.60% (0.55%, 0.71%) | 18/9/2020 | 5/10/2020 |
| 6 | 160,175 | 1,732 | 1.08% (1.03%, 1.13%) | 1.30% (1.21%, 1.39%) | 16/10/2020 | 2/11/2020 |
| 7 | 168,181 | 1,299 | 0.77% (0.73%, 0.82%) | 0.94% (0.87%, 1.01%) | 13/11/2020 | 3/12/2020 |
| 8 | 167,642 | 2,282 | 1.36% (1.31%, 1.42%) | 1.57% (1.49%, 1.66%) | 06/01/2021* | 22/01/2021 |
| 9 | 165,456 | 689 | 0.42% (0.39%, 0.45%) | 0.49% (0.44%, 0.55%) | 4/2/2021 | 23/2/2021 |
| 10 | 140,844 | 227 | 0.16% (0.14%, 0.18%) | 0.20% (0.17%, 0.23%) | 11/03/2021 | 30/3/2021 |
| 11 | 127,408 | 115 | 0.09% (0.07%, 0.11%) | 0.10% (0.08%, 0.13%) | 15/04/2021 | 03/05/2021 |

\* Includes small number of samples from previous days

Using a constant growth rate model, on average for England, we found evidence for a decline over the period of round 10 to 11 (Table 2, Figure 1) with an estimated R of 0.90 (0.87, 0.94). Fitting a P-spline model to the data we found some suggestion that the rate of decline may have slowed into round 11. However, there was considerable uncertainty about the within-round trend with 33% probability of R > 1 averaged across the whole round.

**Table 2.** Estimates of national growth rates, doubling times and reproduction numbers for round 10 to round 11, and within round 11

| Round | Outcome | Growth rate | R | Prob.<br>R>1 | Growth(+) / Decay(-)<br>rate |
| --- | --- | --- | --- | --- | --- |
| 10 and 11 | All positives | -0.016 ( -0.022 , -0.010 ) | 0.90 ( 0.87 , 0.94 ) | <0.01 | -44.2 ( -31.7 , * ) |
|  | Non-symptomatics | -0.011 ( -0.020 , -0.003 ) | 0.93 ( 0.88 , 0.98 ) | <0.01 | * ( -35.0 , * ) |
|  | Positive for both<br>E and N genes | -0.012 ( -0.020 , -0.003 ) | 0.93 ( 0.87 , 0.98 ) | <0.01 | * ( -34.0 , * ) |
|  | Positive for both<br>E and N genes or<br>positive only for N<br>gene with CT 35 or<br>less | -0.019 ( -0.026 , -0.013 ) | 0.88 ( 0.84 , 0.92 ) | <0.01 | -35.8 ( -26.3 , * ) |
| 11 | All positives | -0.010 ( -0.054 , 0.034 ) | 0.94 ( 0.69 , 1.23 ) | 0.33 | * ( -12.8 , 20.3 ) |
|  | Non-symptomatics | 0.006 ( -0.050 , 0.062 ) | 1.04 ( 0.71 , 1.44 ) | 0.59 | * ( -13.8 , 11.2 ) |
|  | Positive for both<br>E and N genes | -0.012 ( -0.073 , 0.046 ) | 0.92 ( 0.60 , 1.32 ) | 0.34 | * ( -9.5 , 15.0 ) |
|  | Positive for both<br>E and N genes or<br>positive only for N<br>gene with CT 35 or<br>less | -0.014 ( -0.067 , 0.036 ) | 0.91 ( 0.62 , 1.25 ) | 0.29 | -48.3 ( -10.4 , 19.1 ) |

\* Doubling/Halving time had an estimated magnitude greater than 50 days and so represented approximately constant prevalence

**Figure 1.**
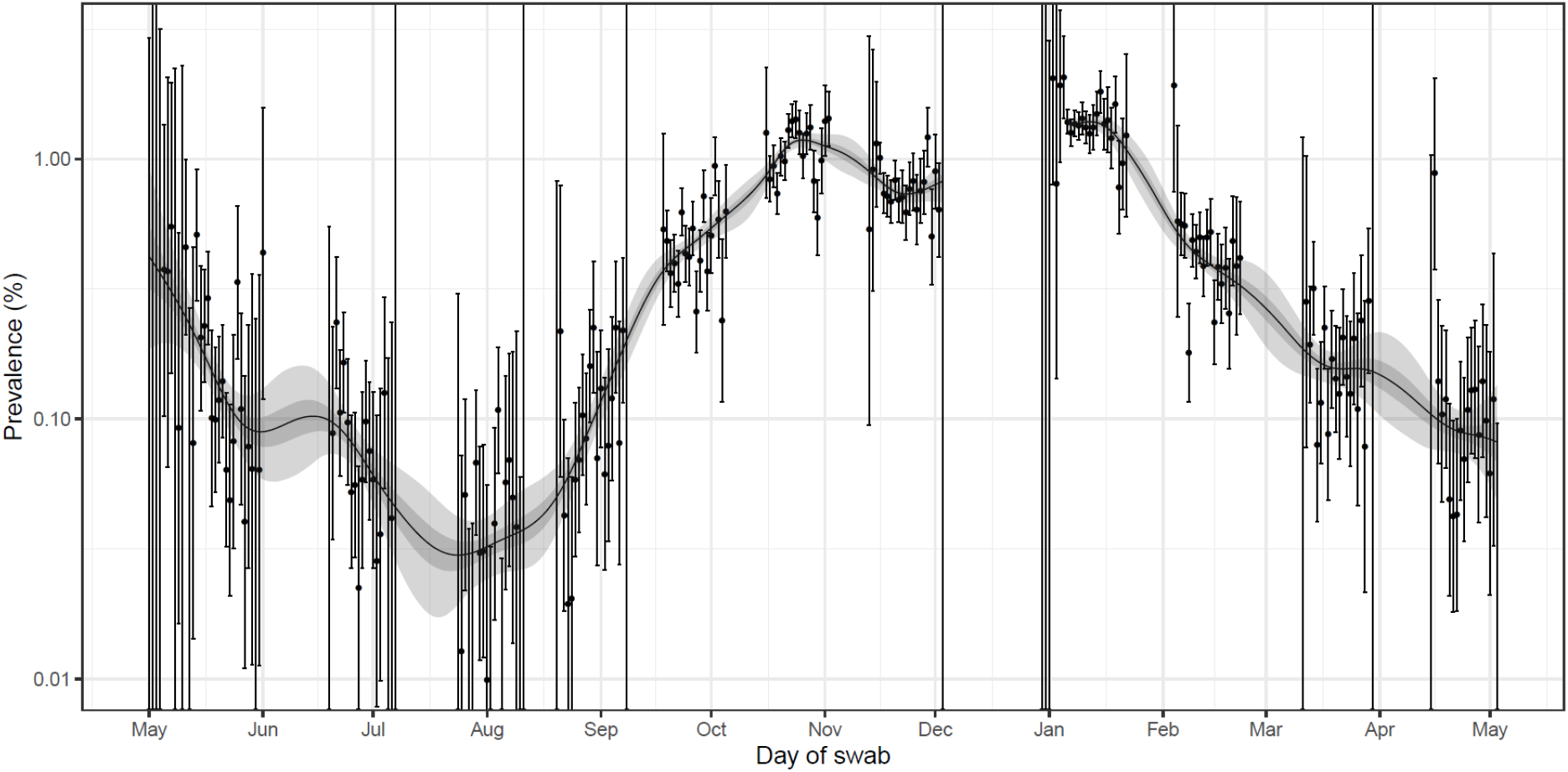
Prevalence of national swab-positivity for England estimated using a P-spline for all eleven rounds with central 50% (dark grey) and 95% (light grey) posterior credible intervals. Shown here only for the entire period of the study with a Log10 y-axis. Unweighted observations (black dots) and 95% binomial confidence intervals (vertical lines) are also shown. Note that the period between round 7 and round 8 (December) of the model is not included as there were no data available to capture the late December peak of the epidemic.

Regional R between rounds 10 to 11 was below one with probability ≥99% in northern regions and East Midlands (Table 3). However, there was a 94% probability of R > 1 in South East. Weighted prevalence across regions (Figure 2, Table 4) ranged from 0.07% (0.03%, 0.17%) in the South West to 0.13% (0.07, 0.27%) in West Midlands.

**Table 3.** Estimates of regional growth rates, doubling times and reproduction numbers for round 10 to round 11.

| Round | Region | Growth rate | R | Prob.<br>R>1 | Halving (-) / Doubling (+) time |
| --- | --- | --- | --- | --- | --- |
| 10 and 11 | East Midlands | -0.019 ( -0.036 , -0.003 ) | 0.88 ( 0.78 , 0.98 ) | 0.01 | -36.4 ( -19.1 , * ) |
|  | West Midlands | -0.015 ( -0.033 , 0.003 ) | 0.91 ( 0.80 , 1.02 ) | 0.05 | -47.4 ( -21.2 , * ) |
|  | East of England | -0.018 ( -0.037 , 0.000 ) | 0.89 ( 0.78 , 1.00 ) | 0.02 | -38.7 ( -18.8 , * ) |
|  | London | -0.013 ( -0.034 , 0.006 ) | 0.92 ( 0.80 , 1.04 ) | 0.08 | * ( -20.6 , * ) |
|  | North West | -0.025 ( -0.042 , -0.009 ) | 0.85 ( 0.75 , 0.94 ) | <0.01 | -27.9 ( -16.7 , * ) |
|  | North East | -0.029 ( -0.058 , -0.004 ) | 0.83 ( 0.67 , 0.97 ) | 0.01 | -23.9 ( -12.0 , * ) |
|  | South East | 0.013 ( -0.003 , 0.030 ) | 1.09 ( 0.98 , 1.20 ) | 0.94 | * ( * , 23.0 ) |
|  | South West | -0.015 ( -0.040 , 0.008 ) | 0.91 ( 0.76 , 1.05 ) | 0.10 | -46.7 ( -17.2 , * ) |
|  | Yorkshire ** | -0.036 ( -0.060 , -0.014 ) | 0.79 ( 0.66 , 0.91 ) | <0.01 | -19.5 ( -11.5 , -48.4 ) |

\* Doubling / halving time had an estimated magnitude greater than 50 days and so represented approximately constant prevalence
\*\* Yorkshire and The Humber

**Table 4a.**
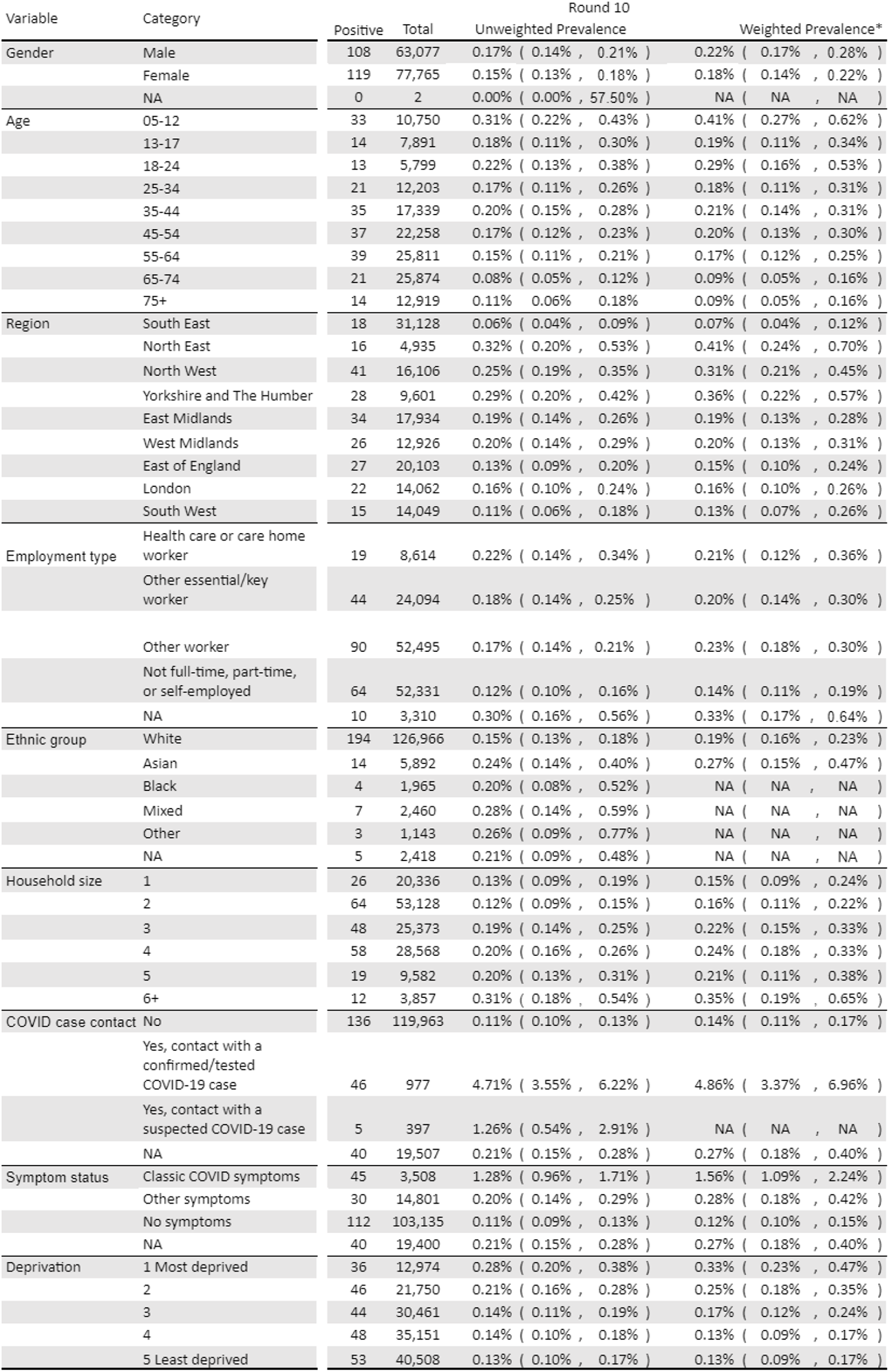
Unweighted and weighted prevalence of swab-positivity for sex age and region for rounds 10. *We present weighted prevalence if the number of positives in a category is 10 or more.

**Table 4b.** Unweighted and weighted prevalence of swab-positivity for sex age and region for rounds 11. *We present weighted prevalence if the number of positives in a category is 10 or more.

| Variable | Category | Round 11 |  |  |  |
| --- | --- | --- | --- | --- | --- |
|  |  | Positive | Total | Unweighted Prevalence | Weighted Prevalence* |
| Gender | Male | 52 | 56,550 | 0.09% ( 0.07% , 0.12% ) | 0.10% ( 0.07% , 0.15% ) |
|  | Female | 63 | 70,855 | 0.09% ( 0.07% , 0.11% ) | 0.11% ( 0.08% , 0.15% ) |
|  | NA | 0 | 3 | 0.00% ( 0.00% , 70.76% ) | NA ( NA , NA ) |
| Age | 05-12 | 12 | 8,551 | 0.14% ( 0.07% , 0.25% ) | 0.16% ( 0.08% , 0.32% ) |
|  | 13-17 | 8 | 6,722 | 0.12% ( 0.05% , 0.23% ) | 0.08% ( 0.04% , 0.17% ) |
|  | 18-24 | 4 | 4,640 | 0.09% ( 0.02% , 0.22% ) | 0.10% ( 0.04% , 0.28% ) |
|  | 25-34 | 17 | 10,238 | 0.17% ( 0.10% , 0.27% ) | 0.21% ( 0.12% , 0.38% ) |
|  | 35-44 | 14 | 14,894 | 0.09% ( 0.05% , 0.16% ) | 0.11% ( 0.05% , 0.21% ) |
|  | 45-54 | 16 | 19,668 | 0.08% ( 0.05% , 0.13% ) | 0.08% ( 0.05% , 0.16% ) |
|  | 55-64 | 18 | 25,330 | 0.07% ( 0.04% , 0.11% ) | 0.06% ( 0.04% , 0.11% ) |
|  | 65-74 | 19 | 24,680 | 0.08% ( 0.05% , 0.12% ) | 0.08% ( 0.05% , 0.13% ) |
|  | 75+ | 7 | 12,685 | 0.06% ( 0.02% , 0.11% ) | 0.05% ( 0.02% , 0.11% ) |
| Region | South East | 26 | 28,262 | 0.09% ( 0.06% , 0.13% ) | 0.11% ( 0.07% , 0.17% ) |
|  | North East | 6 | 4,654 | 0.13% ( 0.05% , 0.28% ) | 0.11% ( 0.05% , 0.25% ) |
|  | North West | 15 | 14,533 | 0.10% ( 0.06% , 0.17% ) | 0.11% ( 0.05% , 0.21% ) |
|  | Yorkshire and The Humber | 6 | 8,629 | 0.07% ( 0.03% , 0.15% ) | 0.12% ( 0.04% , 0.33% ) |
|  | East Midlands | 15 | 16,224 | 0.09% ( 0.05% , 0.15% ) | 0.08% ( 0.05% , 0.14% ) |
|  | West Midlands | 15 | 11,868 | 0.13% ( 0.07% , 0.21% ) | 0.13% ( 0.07% , 0.27% ) |
|  | East of England | 13 | 18,262 | 0.07% ( 0.04% , 0.12% ) | 0.08% ( 0.04% , 0.14% ) |
|  | London | 11 | 12,543 | 0.09% ( 0.04% , 0.16% ) | 0.12% ( 0.06% , 0.24% ) |
|  | South West | 8 | 12,433 | 0.06% ( 0.03% , 0.13% ) | 0.07% ( 0.03% , 0.17% ) |
| Employment type | Health care or care home worker | 6 | 8,384 | 0.07% ( 0.03% , 0.16% ) | NA ( NA , NA ) |
|  | Other essential/key worker | 26 | 20,321 | 0.13% ( 0.08% , 0.19% ) | 0.12% ( 0.08% , 0.19% ) |
|  | Other worker | 43 | 46,761 | 0.09% ( 0.07% , 0.12% ) | 0.11% ( 0.07% , 0.17% ) |
|  | Not full-time, part-time, or self-employed | 36 | 49,601 | 0.07% ( 0.05% , 0.10% ) | 0.08% ( 0.05% , 0.12% ) |
|  | NA | 4 | 2,341 | 0.17% ( 0.05% , 0.44% ) | NA ( NA , NA ) |
| Ethnic group | White | 98 | 114,853 | 0.09% ( 0.07% , 0.10% ) | 0.09% ( 0.07% , 0.11% ) |
|  | Asian | 12 | 5,400 | 0.22% ( 0.11% , 0.39% ) | 0.31% ( 0.16% , 0.60% ) |
|  | Black | 2 | 1,752 | 0.11% ( 0.01% , 0.41% ) | NA ( NA , NA ) |
|  | Mixed | 0 | 2,086 | 0.00% ( 0.00% , 0.18% ) | NA ( NA , NA ) |
|  | Other | 2 | 1,036 | 0.19% ( 0.02% , 0.70% ) | NA ( NA , NA ) |
|  | NA | 1 | 2,281 | 0.04% ( 0.00% , 0.24% ) | NA ( NA , NA ) |
| Household size | 1 | 16 | 19,394 | 0.08% ( 0.05% , 0.13% ) | 0.07% ( 0.04% , 0.14% ) |
|  | 2 | 27 | 49,884 | 0.05% ( 0.04% , 0.08% ) | 0.06% ( 0.04% , 0.09% ) |
|  | 3 | 25 | 22,085 | 0.11% ( 0.07% , 0.17% ) | 0.12% ( 0.07% , 0.22% ) |
|  | 4 | 35 | 24,562 | 0.14% ( 0.10% , 0.20% ) | 0.17% ( 0.11% , 0.26% ) |
|  | 5 | 6 | 8,196 | 0.07% ( 0.03% , 0.16% ) | NA ( NA , NA ) |
|  | 6+ | 6 | 3,287 | 0.18% ( 0.07% , 0.40% ) | NA ( NA , NA ) |
| COVID case contact | No | 71 | 110,070 | 0.06% ( 0.05% , 0.08% ) | 0.07% ( 0.05% , 0.09% ) |
|  | Yes, contact with a confirmed/tested COVID-19 case | 22 | 383 | 5.74% ( 3.63% , 8.57% ) | 4.64% ( 2.77% , 7.67% ) |
|  | Yes, contact with a suspected COVID-19 case | 1 | 198 | 0.51% ( 0.01% , 2.78% ) | NA ( NA , NA ) |
|  | NA | 21 | 16,757 | 0.13% ( 0.08% , 0.19% ) | 0.19% ( 0.11% , 0.34% ) |
| Symptom status | Classic COVID symptoms | 14 | 2,677 | 0.52% ( 0.29% , 0.88% ) | 0.79% ( 0.38% , 1.66% ) |
|  | Other symptoms | 12 | 12,394 | 0.10% ( 0.05% , 0.17% ) | 0.09% ( 0.05% , 0.17% ) |
|  | No symptoms | 69 | 95,642 | 0.07% ( 0.06% , 0.09% ) | 0.07% ( 0.05% , 0.10% ) |
|  | NA | 20 | 16,695 | 0.12% ( 0.07% , 0.19% ) | 0.16% ( 0.09% , 0.28% ) |
| Deprivation | 1 Most deprived | 14 | 11,599 | 0.12% ( 0.07% , 0.20% ) | 0.15% ( 0.08% , 0.28% ) |
|  | 2 | 25 | 19,323 | 0.13% ( 0.08% , 0.19% ) | 0.14% ( 0.09% , 0.22% ) |
|  | 3 | 14 | 27,056 | 0.05% ( 0.03% , 0.09% ) | 0.06% ( 0.03% , 0.11% ) |
|  | 4 | 34 | 32,105 | 0.11% ( 0.07% , 0.15% ) | 0.12% ( 0.08% , 0.18% ) |
|  | 5 Least deprived | 28 | 37,325 | 0.08% ( 0.05% , 0.11% ) | 0.07% ( 0.04% , 0.10% ) |

**Figure 2.**
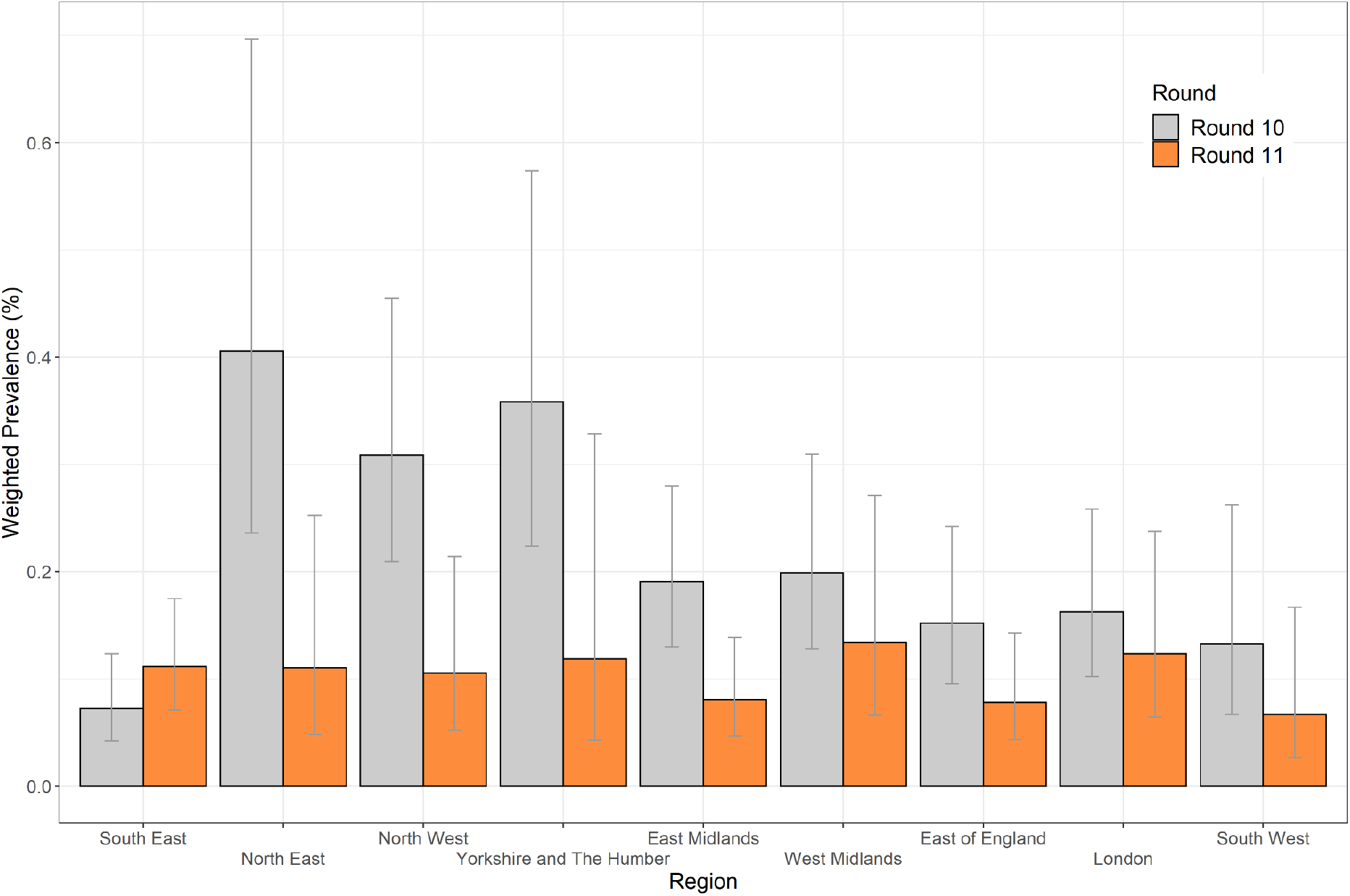
Weighted prevalence of swab-positivity by region for rounds 10 and 11. Bars show 95% confidence intervals.

Weighted prevalence has fallen in 55 to 64 year olds from 0.17% (0.12%, 0.25%) in round 10 to 0.06% (0.04%, 0.11%) in round 11 (Figure 3, Table 4). Weighted prevalence was highest in 25-34 year olds at 0.21% (0.12, 0.38%) similar to the prevalence of 0.18% (0.11%, 0.31%) in round 10. Weighted prevalence amongst participants of Asian ethnicity in round 11 was higher at 0.31% (0.16%, 0.60%) compared with white participants at 0.09% (0.07%, 0.11%) (Table 4). In multivariable logistic regression, with adjustment for core variables, the odds ratio for participants of Asian ethnicity versus white participants was 1.88 (0.95, 3.74) (Table 5). When the analysis was restricted to positive samples with N-gene Ct values less than 34, the odds ratio was 2.47 (1.13, 5.39).

**Table 5.** Multivariable logistic regression for round 10 and round 11.

| Variable | Category | Round 10 | Round 11 | Round 11<br>(N-gene CT < 34)* |
| --- | --- | --- | --- | --- |
| Sex | Male | Ref | Ref | Ref |
|  | Female | 0.87 [0.66,1.15] | 1.02 [0.69,1.49] | 1.15 [0.71,1.84] |
| Age Group | 5-12 | 1.30 [0.79,2.13] | 1.52 [0.69,3.36] | 1.35 [0.50,3.66] |
|  | 13-17 | 0.78 [0.37,1.65] | 1.66 [0.61,4.50] | 1.65 [0.50,5.38] |
|  | 18-24 | 0.82 [0.40,1.67] | 1.03 [0.33,3.19] | 1.48 [0.45,4.87] |
|  | 25-34 | 0.82 [0.47,1.42] | 2.02 [0.97,4.22] | 1.98 [0.82,4.81] |
|  | 35-44 | Ref | Ref | Ref |
|  | 45-54 | 0.81 [0.51,1.30] | 1.01 [0.48,2.10] | 0.74 [0.28,1.92] |
|  | 55-64 | 0.81 [0.49,1.32] | 1.09 [0.52,2.31] | 0.92 [0.36,2.34] |
|  | 65+ | 0.54 [0.31,0.96] | 1.49 [0.66,3.33] | 1.43 [0.54,3.77] |
| Region | North East | 5.65 [2.82,11.30] | 1.38 [0.56,3.42] | 1.01 [0.29,3.51] |
|  | North West | 4.06 [2.27,7.27] | 1.10 [0.57,2.12] | 1.08 [0.48,2.41] |
|  | Yorkshire and The Humber | 4.80 [2.58,8.94] | 0.79 [0.32,1.95] | 0.60 [0.17,2.05] |
|  | East Midlands | 3.29 [1.82,5.94] | 0.96 [0.50,1.86] | 0.92 [0.41,2.07] |
|  | West Midlands | 3.21 [1.71,6.02] | 1.38 [0.72,2.63] | 1.63 [0.77,3.45] |
|  | East of England | 2.36 [1.28,4.36] | 0.69 [0.34,1.41] | 0.65 [0.27,1.56] |
|  | London | 2.20 [1.14,4.25] | 0.71 [0.33,1.53] | 0.67 [0.27,1.69] |
|  | South East | Ref | Ref | Ref |
|  | South West | 2.02 [1.01,4.06] | 0.76 [0.34,1.69] | 0.57 [0.19,1.71] |
| Key Worker Status | HCW/CHW | 0.99 [0.58,1.68] | 0.70 [0.29,1.66] | 0.49 [0.15,1.62] |
|  | Key worker (other) | 0.91 [0.63,1.31] | 1.31 [0.79,2.15] | 1.03 [0.54,1.98] |
|  | Other worker | Ref | Ref | Ref |
|  | Not FT, PT, SE | 0.88 [0.60,1.27] | 0.78 [0.46,1.31] | 0.76 [0.41,1.41] |
| Ethnicity | White | Ref | Ref | Ref |
|  | Asian | 1.32 [0.74,2.37] | 1.95 [0.98,3.88] | 2.47 [1.13,5.39] |
|  | Black | 1.11 [0.40,3.07] | 1.26 [0.30,5.25] | 2.04 [0.48,8.70] |
|  | Mixed | 1.69 [0.78,3.66] | 0.00 [0.00,2.821236e+136] | 0.00 [0.00,3.773294e+167] |
|  | Other | 1.65 [0.52,5.26] | 2.24 [0.54,9.27] | 1.77 [0.24,13.02] |
| Household Size | 1-2 People | Ref | Ref | Ref |
|  | 3-5 People | 1.27 [0.90,1.78] | 1.84 [1.13,2.98] | 1.71 [0.94,3.12] |
|  | 6+ People | 1.42 [0.68,2.97] | 2.19 [0.81,5.94] | 1.12 [0.25,5.05] |
| Deprivation Index Quintile | 1 - Most Deprived | Ref | Ref | Ref |
|  | 2 | 0.94 [0.60,1.50] | 1.03 [0.52,2.03] | 1.05 [0.47,2.35] |
|  | 3 | 0.69 [0.43,1.11] | 0.50 [0.23,1.06] | 0.45 [0.18,1.14] |
|  | 4 | 0.66 [0.42,1.05] | 0.98 [0.51,1.86] | 0.71 [0.32,1.60] |
|  | 5 - Least Deprived | 0.68 [0.43,1.08] | 0.68 [0.35,1.34] | 0.71 [0.32,1.58] |

\* CT cutoff of 34 selects the ~1/2 positive samples with highest viral load

**Figure 3.**
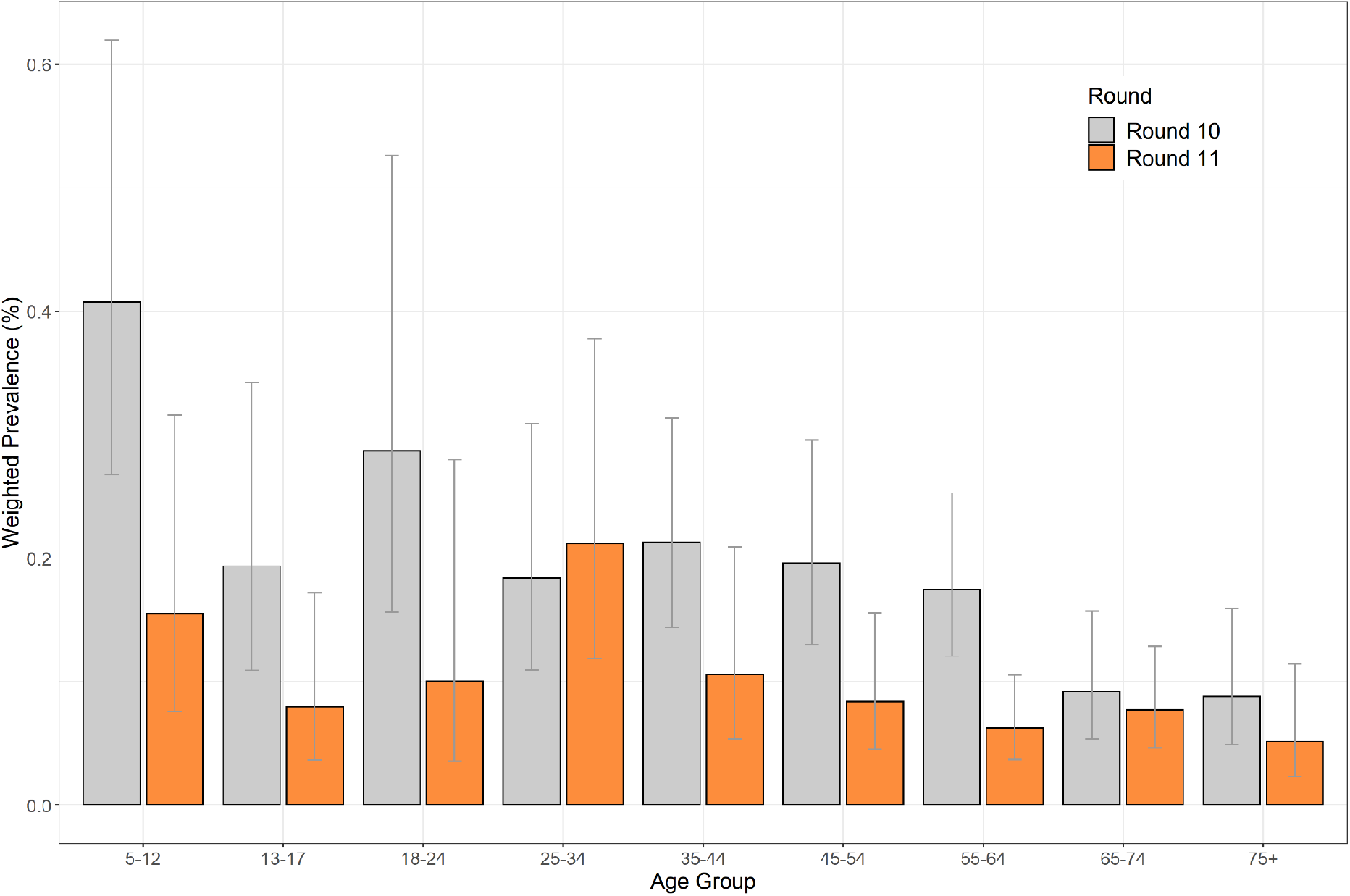
Weighted prevalence of swab-positivity by age for rounds 10 and 11. Bars show 95% confidence intervals.

In round 11, mean N- and E-gene Ct values were higher relative to round 8 (January), 9 (February) and 10 (March) (Table 6). After restricting analysis to positive samples in which both N- and E-gene were detected, we found a mean N-gene Ct value of 29.1 (27.7, 30.5) in round 11 compared to 24.8 (24.6, 25.1) in round 8 (*P* <0.001), and a mean E-gene Ct value of 29.1 (27.6, 30.7) in round 11 compared to 26.5 (26.2, 26.8) in round 8 (*P*<0.001).

**Table 6.**
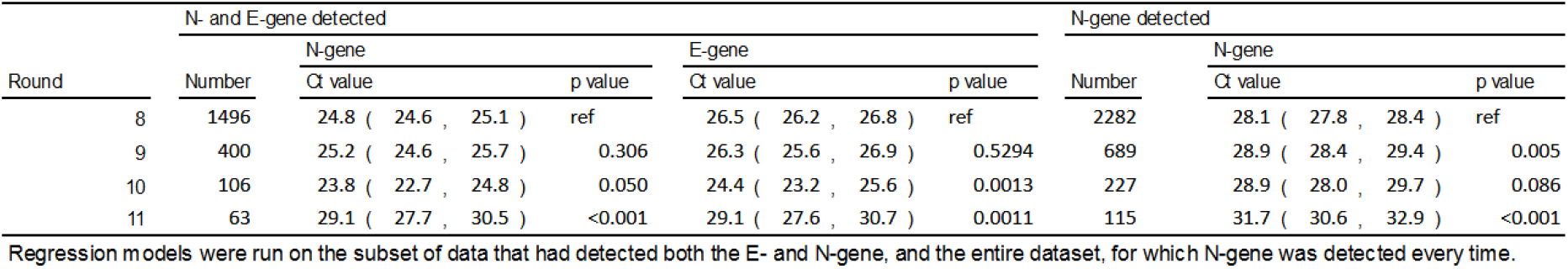
Results of Gaussian regression with either E-gene or N-gene Ct value as the observation and REACT-1 round as the explanatory variable.

For round 11, lineages have been determined for 26 of the 115 positive swab tests obtained. We identified B.1.1.7 and B.1.617.2 lineages: 92.3% (75.9%, 97.9%, n=24) B.1.1.7 and 7.7% (2.1%, 24.1%, n=2) B.1.617.2. Both samples from the B.1.617.2 lineage were detected in London (out of three positives overall for London for which a lineage was determined) in people from different local areas who did not report returning from abroad in the previous two weeks.

We observed a divergence from the prior close alignment between incidence measured in REACT-1 and hospitalisations and deaths recorded by routine surveillance (with appropriate lag periods, Figure 4, Figure 5).

**Figure 4.**
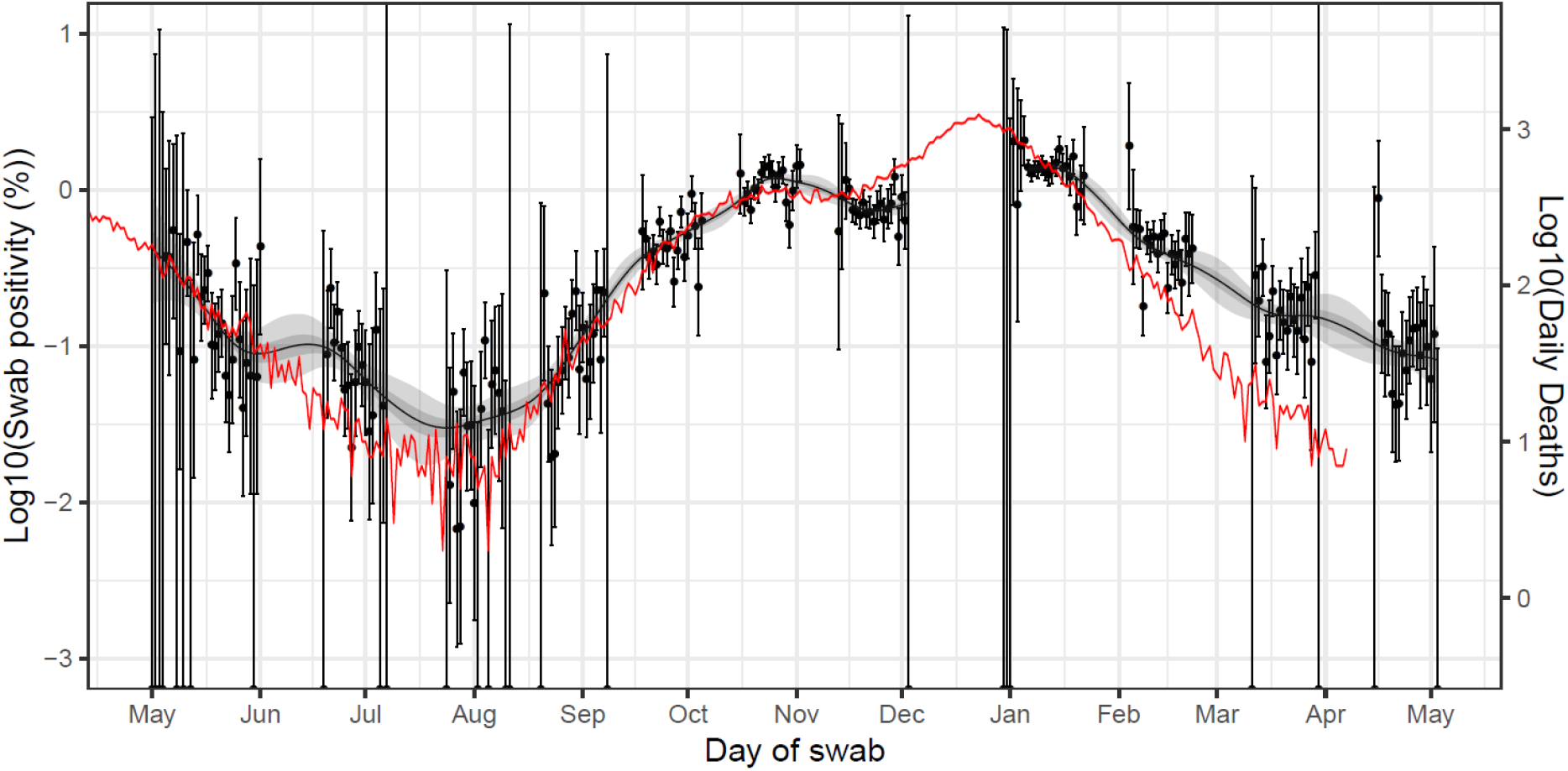
Daily deaths in England (Solid red line, right hand y-axis) shifted by a lag parameter along the x-axis (see below), and daily swab positivity for all 11 rounds of the study (dots with 95% confidence intervals, left hand y-axis) and the P-spline estimate for swab positivity (Solid black line, dark grey shaded area is 50% central credible interval, light grey shaded area is 95% central credible interval, left-hand y-axis). Daily deaths have been fit to observations from the first 10 rounds of the REACT-1 study to obtain scaling and lag parameters. These parameter values were estimated using a Bayesian MCMC model: daily_positives(t) ∼ Binomial(daily_swab_tests(t), p = daily_admissions(t+lag)*scale). The time lag parameter was estimated at 27 (27, 27) days. Note the P-spline is not plotted for the region between round 7 and 8 in which there was an unobserved peak in swab-positivity.

**Figure 5.**
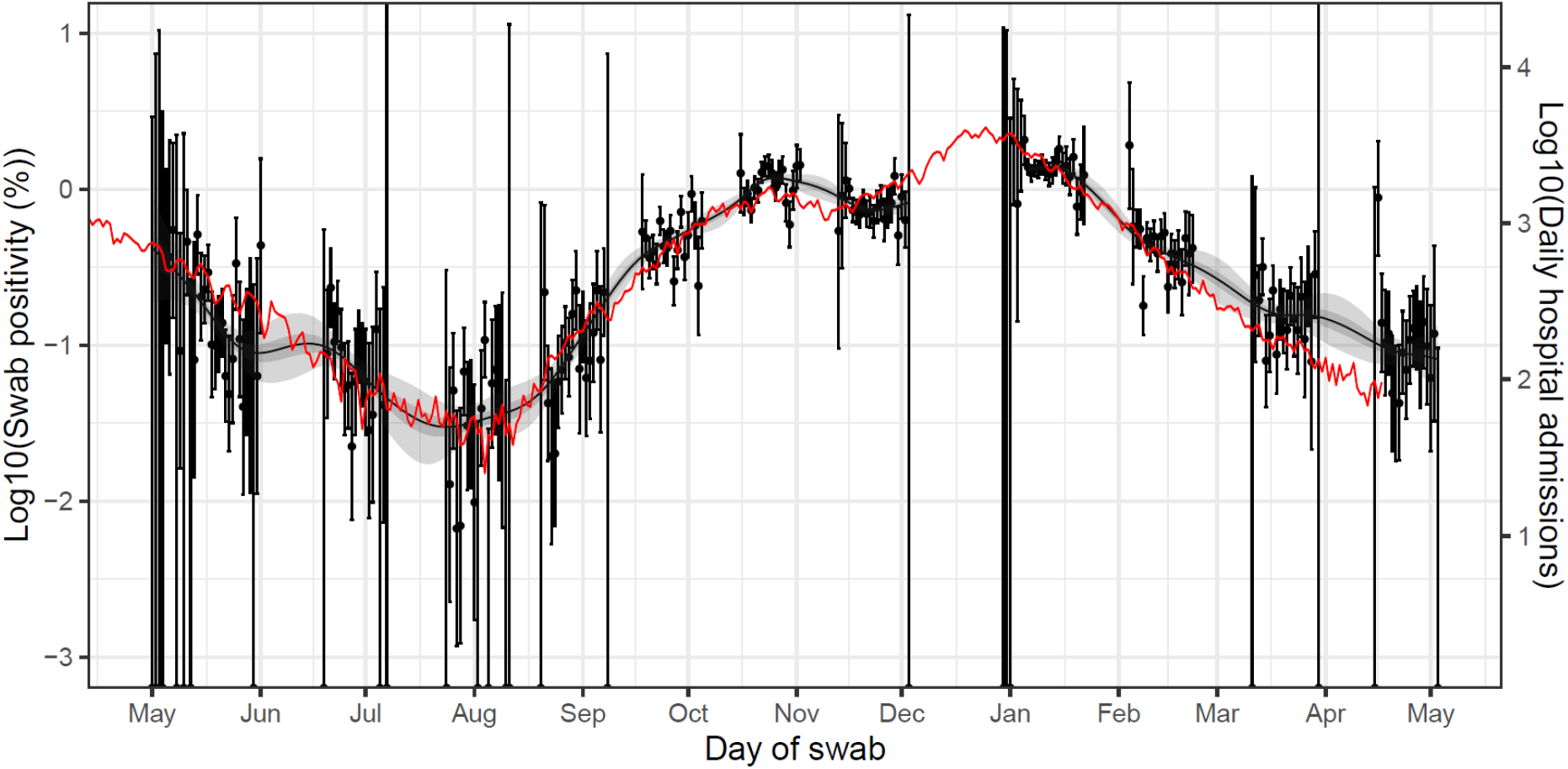
Daily hospital admissions in England (Solid red line, right hand y-axis) shifted by a lag parameter along the x-axis (see below), and daily swab positivity for all 11 rounds of the study (dots with 95% confidence intervals, left hand y-axis) and the P-spline estimate for swab positivity (Solid black line, dark grey shaded area is 50% central credible interval, light grey shaded area is 95% central credible interval, left-hand y-axis). Daily hospital admissions have been fit to observations from the first 10 rounds of the REACT-1 study to obtain scaling and lag parameters. These parameter values were estimated using a Bayesian MCMC model: daily_positives(t) ∼ Binomial(daily_swab_tests(t), p = daily_admissions(t+lag)*scale). The time lag parameter was estimated at 18 (16, 18) days. Note the P-spline is not plotted for the region between round 7 and 8 in which there was an unobserved peak in swab-positivity.

## Discussion

In this eleventh round of sampling in the REACT-1 study we observed a decline in prevalence of 50% in comparison with the previous round (March 2021). This period was during the third national lockdown in England, which began in early January 2021 following a rapid rise of infections during November-December 2020 [8]. This rapid increase in infections coincided with the spread of the B.1.1.7 lineage, first observed in Kent, England, in September 2020, which then became the dominant lineage in England during February and March 2021 [9].

Between rounds 10 and 11 there was a two-thirds fall in prevalence among those aged 55 to 64 years old, which may reflect the effects of the recent roll-out of the vaccination programme in this age group. Evidence from the vaccination programme in Israel (two doses of the BNT162b2 mRNA vaccine [10]) suggests a 95% efficacy against infection [11]. On the other hand, we observed similar rates in rounds 10 and 11 for 25 to 34 year olds against a background of reducing prevalence, which may reflect increased contact rates, due to the easing of restrictions, in this age group where there are low levels of vaccination coverage.

Round 11 Ct values were higher than those in round 10, corresponding to a reduction in viral load and suggesting fewer recent infections [12]. Prevalence rates are now at levels last seen in summer 2020; compared with periods of peak prevalence, at these levels, a higher proportion of positive cases detected by PCR may be from people who continue to shed the virus over prolonged periods rather than from lower-Ct recent infections [13].

We saw higher prevalence in people of Asian ethnicity compared with white people, especially if we restricted the analyses to Ct <34, that is, where viral load was higher. This suggests that infections may be continuing to spread more rapidly among the Asian community than in other groups in the population.

Our viral sequencing data indicate that B.1.1.7 lineage remains dominant in England (92% of sequenced samples) but that B.1.617.2 is also circulating in London, consistent with reports from Public Health England (PHE) [14]. According to PHE, only 22% of B.1.617.2 cases reported to date were associated with international travel; the two cases of B.1.617.2 identified in our study were both among people who had not travelled abroad in the previous two weeks. The fact that we observed B.1.617.2 at a similar (or higher) frequency to the long-established B.1.1.7 lineage in London suggests that B.1.617.2 may be more transmissible than B.1.1.7 in the populations where the two viruses are currently circulating. However further studies are needed to investigate the geographic extent of cases and the degree to which they are explained by one or more of: increased intrinsic transmissibility, variations in social mixing patterns, and differing levels of vaccination. Also, studies are ongoing to measure any possible antigenic difference between B.1.617.2 and B.1.1.7 which could affect vaccine efficacy [14].

Our analyses show divergence in rates between infections as measured in REACT-1 and hospitalisation and mortality trends based on national data. The divergence began in January 2021, coinciding with the mass roll out of the vaccination programme in England to the elderly and most vulnerable in society. This divergence in trends suggests that the vaccination programme may be contributing to lower rates of severe outcomes (hospitalisations and deaths) in the older age groups, effectively breaking the link between infection, hospitalisations and deaths. Our observations are consistent with studies in Scotland [7] and Israel [8] which reported high efficacy of vaccination against severe disease. It will be important to continue to monitor trends in the relationship between these three outcomes to see whether these curves re-converge, which might give an early indication of circulating lineages associated with reduced efficacy of the vaccine.

Our study has limitations. Our sampling may not be fully representative of the population of England, despite attempts to correct for this using weighting. There is now widespread availability of testing in England, including for non-symptomatic people through either ‘surge’ testing in areas of high prevalence or where there are variants of concerns (VOCs), or on demand using lateral flow tests. We have seen a moderate fall in response rate during the course of the study (currently 15% of people to whom we send invite letters provide a viable swab), which may reflect lower levels of public concern in recent weeks. Nonetheless, we believe that our estimates of prevalence and R are broadly representative of the population, and likely less affected by these changes in testing behaviour than routine surveillance data [6]. Reliable sequencing data were only obtained for 20% of the positive samples, since at higher Ct values (when there is less virus present) good sequence coverage is difficult to obtain.

If prevalence rises in the coming weeks, policy-makers will need to assess the possible impact on hospitalisations and deaths. In addition, consideration should be given to other health and economic impacts if increased levels of community transmission occur.

## Methods

In REACT-1 we collect a self-administered throat and nose swab sample and questionnaire data from a random sample of the population in England at ages 5 years and above (parent/guardian administered at ages 5 to 12) [7]. We use the National Health Service (NHS) register of patients to select the sample aiming to obtain similar numbers of participants in each of the 315 lower tier local authorities (LTLAs) in England. Completed swabs are placed in the home refrigerator before being picked up by courier and sent chilled to the laboratory for RT-PCR testing. We sent out 823,812 invitations to people registered with a GP on the NHS register and dispatched 107,503 (20.7%) test kits. This resulted in 127,408 (74.7%) completed swabs with a valid test result, giving an overall response rate of 15.5% (valid swabs divided by total number of invites).

We estimate prevalence of RT-PCR swab-positivity with and without weighting, both to allow for the sample design (balanced by LTLA, not by population) and to adjust for variable non-response. The aim is to provide prevalence estimates that are representative of the population of England as a whole, by age, sex, region, ethnicity and other socio-demographic characteristics.

We estimate the reproduction number R using exponential growth models, both between successive rounds and within rounds. In sensitivity analyses, we provide estimates of R for i) different cut-points of cycle threshold (Ct) values for swab-positivity and ii) after restricting the analyses to those not reporting symptoms in the previous week.

We fit a smoothed P-spline function to the daily prevalence data across all rounds, with knots at 5-day intervals, to examine trends in unweighted prevalence over time [15]. Using an appropriate scaling parameter and suitable discrete-day lag periods, we compare the daily data on prevalence in REACT-1 with the publicly available national daily hospital admissions and COVID-19 mortality data (deaths within 28 days of a positive test), to examine whether there have been any changes in the relationships of prevalence to hospitalisations and deaths.

We carried out genome sequencing using the Illumina NextSeq 500 platform for PCR positive swab samples where there was sufficient sample volume and with N-gene Ct values < 32. Viral RNA was amplified using the ARTIC protocol [16] with sequencing libraries prepared using CoronaHiT [17]. Sequencing data were analyzed using the ARTIC bioinformatic pipeline [18] with lineages assigned using PangoLEARN [19].

We carried out statistical analyses in R [14]. Research ethics approval was obtained from the South Central-Berkshire B Research Ethics Committee (IRAS ID: 283787). The COG-UK study protocol was approved by the Public Health England Research Ethics Governance Group (reference: R&D NR0195).

## Data Availability

Links to a spreadsheet, and the GitHub R pacakge containing supporting data for tables and figures are available within the manuscript.

## Data availability

Supporting data for tables and figures are available either: in this [spreadsheet](https://docs.google.com/spreadsheets/d/1QJi4h4JWWP_hSaGDdcMSQ1RxgzM2SSo9tV-4ookIHjk/edit#gid=1065915288); or in the inst/extdata directory of this GitHub R package. Assembled/consensus genomes are available from GISAID subject to minimum quality control criteria. Raw reads are available from European Nucleotide Archive (ENA). All genomes, phylogenetic trees, and basic metadata are available from the COG-UK consortium website (https://www.cogconsortium.uk).

## Acknowledgements

SR, CAD acknowledge support: MRC Centre for Global Infectious Disease Analysis, National Institute for Health Research (NIHR) Health Protection Research Unit (HPRU), Wellcome Trust (200861/Z/16/Z, 200187/Z/15/Z), and Centres for Disease Control and Prevention (US, U01CK0005-01-02). NFA was supported by the Quadram Institute Bioscience BBSRC funded Core Capability Grant (project number BB/CCG1860/1). GC is supported by an NIHR Professorship. HW acknowledges support from an NIHR Senior Investigator Award and the Wellcome Trust (205456/Z/16/Z). PE is Director of the MRC Centre for Environment and Health (MR/L01341X/1, MR/S019669/1). PE acknowledges support from Health Data Research UK (HDR UK); the NIHR Imperial Biomedical Research Centre; NIHR HPRUs in Chemical and Radiation Threats and Hazards, and Environmental Exposures and Health; the British Heart Foundation Centre for Research Excellence at Imperial College London (RE/18/4/34215); and the UK Dementia Research Institute at Imperial (MC_PC_17114). We thank The Huo Family Foundation for their support of our work on COVID-19. Quadram authors gratefully acknowledge the support of the Biotechnology and Biological Sciences Research Council (BBSRC); their research was funded by the BBSRC Institute Strategic Programme Microbes in the Food Chain BB/R012504/1 and its constituent project BBS/E/F/000PR10352. We thank members of the COVID-19 Genomics Consortium UK for their contributions to generating the genomic data used in this study. The COVID-19 Genomics UK (COG-UK) Consortium is supported by funding from the Medical Research Council (MRC) part of UK Research & Innovation (UKRI), the National Institute of Health Research (NIHR) and Genome Research Limited, operating as the Wellcome Sanger Institute.

We thank key collaborators on this work – Ipsos MORI: Kelly Beaver, Sam Clemens, Gary Welch, Nicholas Gilby, Kelly Ward and Kevin Pickering; Institute of Global Health Innovation at Imperial College: Gianluca Fontana, Sutha Satkunarajah, Didi Thompson and Lenny Naar; Molecular Diagnostic Unit, Imperial College London: Prof. Graham Taylor; North West London Pathology and Public Health England for help in calibration of the laboratory analyses; Patient Experience Research Centre at Imperial College and the REACT Public Advisory Panel; NHS Digital for access to the NHS register; and the Department of Health and Social Care for logistic support. SR acknowledges helpful discussion with attendees of meetings of the UK Government Office for Science (GO-Science) Scientific Pandemic Influenza – Modelling (SPI-M) committee.

## Additional information

Full list of COG-UK author’s names and affiliations are available in this [spreadsheet](https://drive.google.com/file/d/11Q3cnp2qGKMqcfrTKpp53jZFc7OdF0f7/view?usp=sharing).

